# Towards a Framework for Case Identification in Pharmacovigilance: Not All Reports are Created Equal

**DOI:** 10.64898/2026.06.23.26354546

**Authors:** M. Fusaroli, J. Félix China, D. Sartori, V. Giunchi, L. Härmark, J. Scholl, F. van Hunsel, G. N. Norén, J. Ellenius

## Abstract

**Background:** Retrieval of adverse event reports based on coded drug-event co-occurrence enables large-scale pharmacovigilance analyses, but yields candidate reports rather than validated cases, risking misinterpretation if used alone.

**Aim:** To develop and apply a framework for identification and characterization of clinically meaningful case series in pharmacovigilance.

**Methods:** We conducted two case studies. The first developed and refined the framework in an information-rich setting, focusing on drug-induced impulsivity across selected drugs; the second tested its applicability in a more routine, information-poor setting, focusing on drug-induced suicidality.

**Results:** In Case 1, non-relevant reports were frequent for drugs with uncertain evidence and negative controls (≈20-40%) compared to drugs with established causal roles (4%). The emerging framework assessed relevance based on exposure, event, drug-event relationship, and population. For suspected adverse drug reactions, relevant reports were further characterized by reporter suspicion and evidentiary qualifiers supporting or refuting causality; higher suspicion was associated with more supportive qualifiers. Applied to Case 2, the framework ruled out 69% of reports as non-relevant but highlighted substantial non-assessability (17%).

**Conclusions:** In pharmacovigilance, retrieval is not equivalent to case identification. Relevance is question-specific and shaped by how reports are captured, processed, and retrieved. This can be especially critical for emerging or bias-prone safety questions. Transparent and reproducible case definition and adjudication are essential for interpretable analyses.

**Graphical Abstract:** 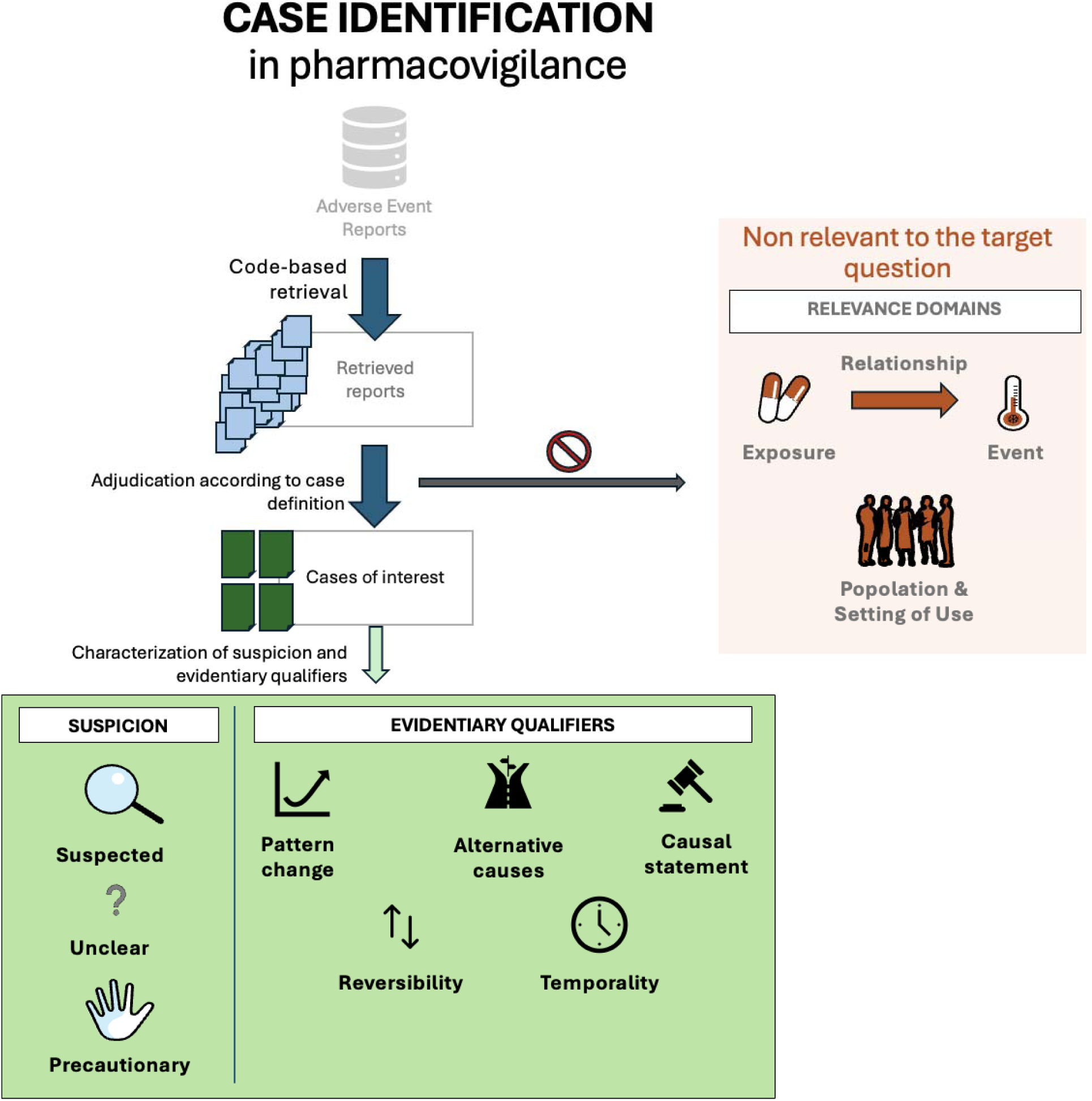

Code-based retrieval of drug–event co-occurrence yields a heterogeneous set of candidate reports that does not directly correspond to a clinically meaningful case series. The proposed framework separates case identification from retrieval by adjudicating relevance across exposure, event, relationship, and context, ruling out non-relevant or non-assessable reports. Relevant reports are further qualified by reporter suspicion and evidentiary features, enabling more interpretable pharmacovigilance analyses.

## 1. Introduction

**Large adverse event reporting systems collect individual case reports of suspected adverse drug reactions for post-marketing safety analysis**, but in practice capture a broader range of medicine-related problems (e.g., lack of effect, medication errors, misuse, product quality issues [1]). Reports combine narrative clinical description with structured data, standardized through formats such as ICH-E2B [2] and coding systems like MedDRA (Medical Dictionary for Regulatory Activities) [3].

**Retrieval based on coded drug-event co-occurrence^1^ enables scalable analyses but yields sets of candidate reports that may not represent a coherent clinical phenomenon**. Structured fields cannot fully preserve narrative meaning, and coding systems lack operational definitions that ensure alignment between reporting, coding, and analytic intent [7]. As a result, coding and processing may introduce information loss, and reports sharing the same coded drug and event may actually refer to extremely different clinical concepts, ultimately pertaining (i.e., being clinically relevant) to separate case series.

**Regulatory guidance emphasizes the need for explicit case definitions** (i.e., criteria to determine whether a retrieved report should be included in a case series [8]). While existing guidelines and efforts focus mainly on diagnostic validity of the outcome [9,10], a report may also be non-relevant because of other reasons, including non-relevant exposure [11]. Epidemiological guidelines recommend operationalizing not only outcomes, but also exposures and covariates in ways that are fit for purpose [12], and integrating both structured and unstructured data [13].

**Without systematic assessment of relevance, reports may be incorrectly assigned** in case series or contingency tables [14], affecting relevance and interpretability of the analytical data output [15] both in qualitative [16] and quantitative analyses [17,18]. Although manual review often addresses this in practice, the distinction between retrieved reports and relevant cases is not always explicit or reproducible.

## 2. Aim

The aim of this study is to **develop and evaluate a preliminary framework to support systematic and transparent identification of** clinically meaningful case series **using two case studies**.

## 3. Methods

### 3.1 Study Design Overview

**We conducted a sequential framework development study** [19] using VigiBase, the WHO global database of adverse event reports for medicines and vaccines (Figure 1).

- **Phase 1 grounded the framework on existing knowledge.**
- **Phase 2 iteratively developed the framework** under information-rich conditions using drug-induced impulsivity (Case study 1), including drugs with established (positive controls), uncertain, and no (negative controls) evidence for causing impulsivity.
- **Phase 3 applied the framework under routine-review conditions** with variable data availability using drug-induced suicidality (Case study 2), explicitly incorporating assessability.

**Figure 1.**
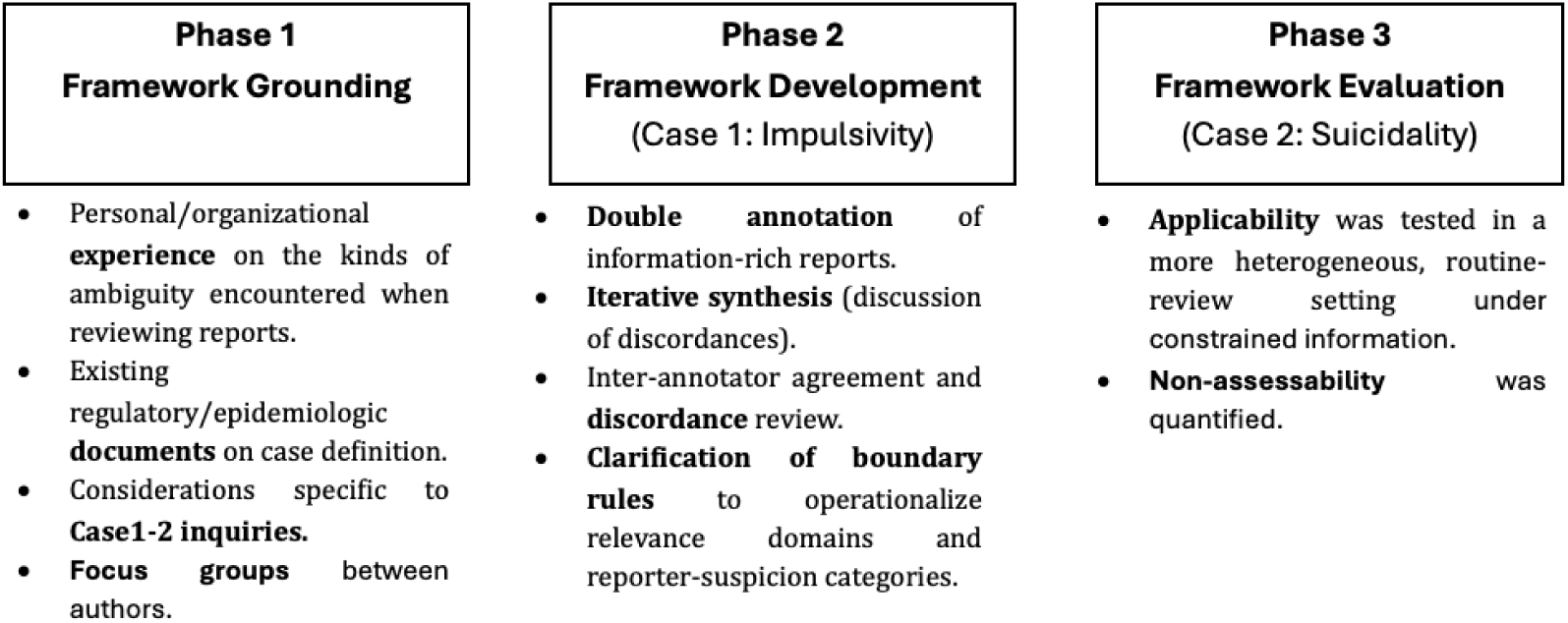
– Study design diagram, showing the three phases of framework development, including sub-steps and reference to the Case studies implemented.

### 3.2 Phase 1: Framework grounding

Regulatory guidance [8,20], prior initiatives (e.g., Brighton Collaboration [9], ENCEPP [21]), MedDRA term-selection guidance^2^ [22], and methodological literature on challenges in coding exposure [11], events [7], and their relationship [1], informed the definition of **four relevance domains**: exposure, event, drug-event relationship, and setting/population (Table 1).

**Table 1.** Relevance domains, grounded in Phase 1 and operationalized in Phase 2. May require adaptation for other clinical questions.

| Relevance Domain | What it captures |
| --- | --- |
| Exposure | • **Misreported or miscoded drug exposure**<br>• **Falsified or substandard products** (potentially coded under the active ingredient they imitate)<br>• **Placebo-arm reports** in blinded trials<br>• **Drug name confusion** (e.g., intended and improperly administered drugs both coded as interacting)<br>• **Non-relevant administration** (e.g., because of non-relevant strength, route, product name, lot, posology, expiration date) |
| Event | • **Misreported event** due to lack of expertise of the reporter<br>• **Miscoded event** due to unclear/non-existent coding recommendations, heterogeneous conventions (e.g., coding individual signs and symptoms, rather than the syndrome, rather than both), or lack of pertinent MedDRA codes.<br>• **Mistranslation** (e.g., *delirio* in Italian should be translated to *delusion* and not to *delirium*)<br>• **Non-verified diagnostic criteria.** |
| Relationship | • **Bystander**: Event explicitly attributed to a different drug (e.g., several drugs suspected for different events in the report, or historical exposures and concomitants improperly coded as suspected)<br>• **Lack of effect** or relapse during treatment<br>• **Reverse causality**: drug taken to treat the event, or because of it (e.g., prophylaxis)<br>• **Discontinuation**: the event occurred after treatment interruption, missed dose, shortage (i.e., when qualifying as withdrawal effect or rebound effect)<br>• **Impossible timing**: the event occurred before exposure (e.g., medical history reported as adverse event). |
| Setting/Population | • **Miscoded demographic data** (e.g., age = 0 can result from incautious data processing)<br>• **Non-relevant care setting**<br>• **Use outside routinary prescription** (e.g., off-label, misuse, abuse, poisoning...) |

Causality principles [23,24] and prior work on report utility (e.g., considering completeness [25,26], evidentiary strength [27,28], reporter suspicion [29]) informed the definition of reporters suspicion and evidentiary qualifiers (Table 2-3), which allows to at least partly capture the different contribution of individual cases in a case series to a question of interest [18].

**Table 2.** Reporter suspicion categories assigned to reports relevant to questions on adverse drug reactions, grounded in Phase 1 and operationalized in Phase 2.

| Suspicion Label | What it captures |
| --- | --- |
| Explicit suspicion | The report explicitly attributed the event to the drug or provided strong supporting evidence. |
| Precautionary | The report explicitly framed the event as plausibly not resulting from the drug. |
| Unclear suspicion | The report did not contain sufficient information to classify it with regard to suspicion. |

**Table 3.** Evidentiary qualifiers identified in reports relevant to questions on adverse drug reactions, grounded in Phase 1 and operationalized in Phase 2. Although these qualifiers may strengthen or weaken suspicion and inform causal reasoning, no combination of qualifiers is sufficient to establish or rule out causality.

| Evidentiary Qualifier | What it captures | Supports suspicion | Weakens suspicion |
| --- | --- | --- | --- |
| **Change in event pattern/severity** | Whether the event newly appears or aggravates. | New onset, escalation, or qualitative change. | No meaningful change from baseline. |
| **Temporality** | Whether the timing is compatible with the suspected drug effect. | The onset followed treatment initiation or a dose change within a plausible timeframe. | Timing was inconsistent with exposure. |
| **Reversibility/ dose relation** | Whether the event changes with withdrawal, rechallenge, or dose modification. | Improved after withdrawal; recurred on rechallenge; worsened with dose increase; improved with dose reduction. | Persisted despite withdrawal; no relation to dose changes. |
| **Alternative explanations** | Whether competing causes are ruled out or favored. | Other explanations were explicitly ruled out. | Another drug or the underlying disease could better explain the event. |
| **Explicit causal statement** | Whether the reporter explicitly affirms or rejects a causal link. | Drug described as causing or likely causing the event. | Drug described as unrelated or unlikely to have caused the event. |

### 3.3 Phase 2: Framework development

**Case study 1** (drug-induced impulsivity) was selected due to overlap with related constructs (e.g., aggressivity, suicidality, compulsivity) and treatment-failure contexts (i.e., impulsivity due to lack of effect). The set up of the study is described in Table 4. Empty and auto-generated narratives were excluded to support framework development under information-rich conditions and reduce annotation burden. Reports were accessed in tabular format (one row per report) to facilitate cross-report comparison and iterative extension and refinement of the annotation.

**Table 4.**
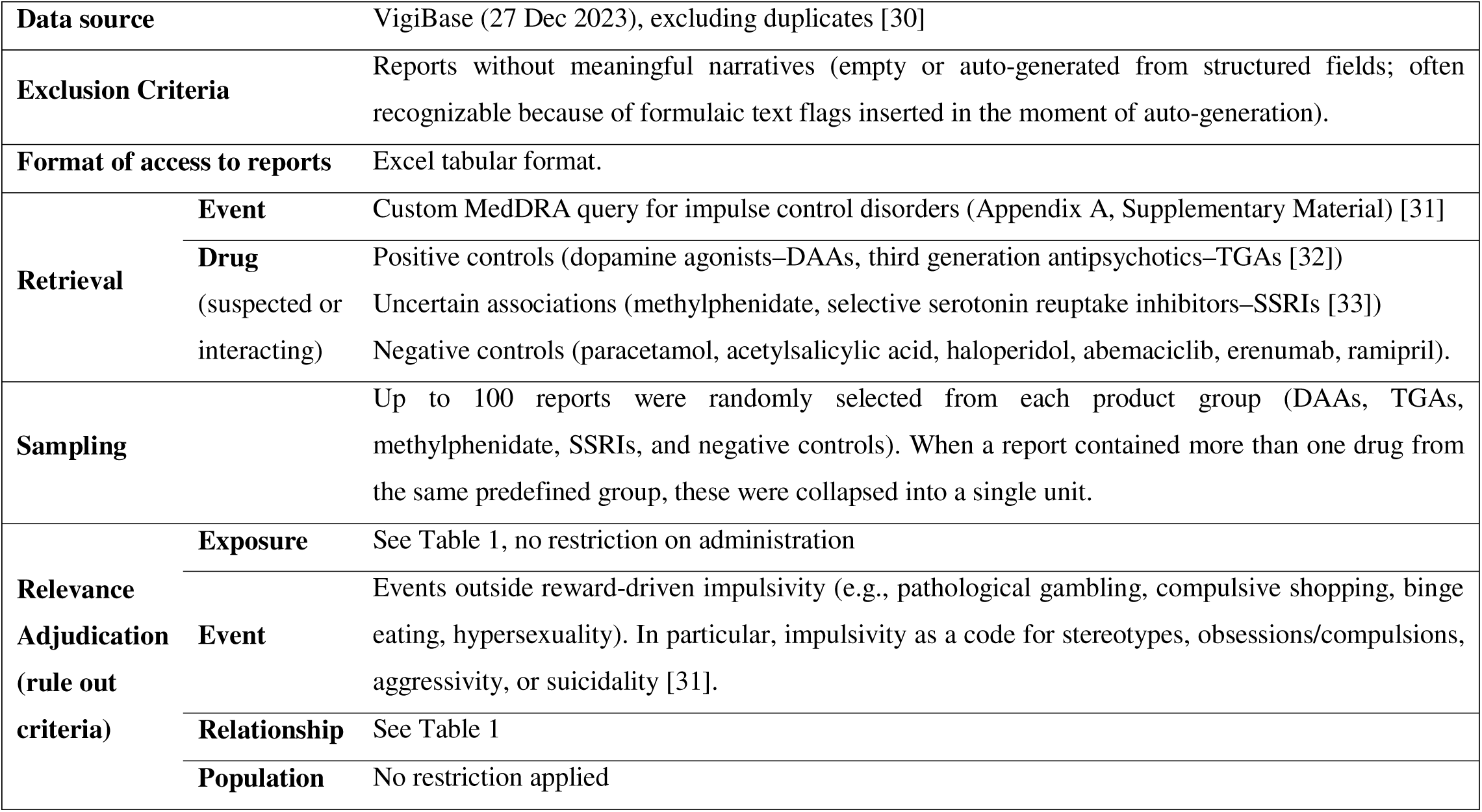
Set up for Case 1, used for the iterative development of the case identification framework.

**Two physicians and pharmacovigilance scientists (co-authors JF, MF) independently annotated** reports at the drug level in relation to the target event construct defined by the specific study question (as different drugs in a report may be more or less relevant to the question; henceforth referred to as report level for simplicity) using structured and unstructured data, assessing relevance, suspicion, and evidentiary qualifiers. The adjudication of suspicion accounted for evidentiary qualifiers (supporting or arguing against a causal relationship), prioritizing explicit causal statements and the patient’s own reported interpretation when available.

**Discordances**, **ambiguous category boundaries, and inconsistency were iteratively discussed to refine decision rules and reach consensus.** Unsolved discordances were excluded (Appendix B, Supplementary Material). Inter-annotator agreement was summarized using Cohen’s k, to assess whether the rules could be applied consistently. Asymmetry in discordant classifications was detected, to identify choices in which the annotators systematically diverged, using the generalized McNemar-Bowker test for paired categorical data [34]. Patterns were compared across product groups to examine whether relevance varied systematically.

### 3.4 Phase 3: Framework evaluation

**Case study 2, on drug-induced suicidality**, was chosen as it is complicated by treatment-failure contexts and overdose as a means of self-harm. The set up of the study is described in Table 5. No exclusions were made based on narrative availability, and individual reports were reviewed in PDF format to better reflect routine case-review conditions (with variable amount of available information) and support richer within report contextualization. When information was insufficient, reports were classified as non-assessable.

**Table 5.** Set up for Case 2, used for testing the applicability of the case identification framework.

|  |  |  |
| --- | --- | --- |
| **Data source** |  | VigiBase (25 Jan 2026), excluding duplicates [35]. |
| **Exclusion Criteria** |  | No exclusions based on narrative availability. |
| **Format of access to reports** |  | PDF format |
| **Retrieval** | **Event** | Narrow Standardized MedDRA Query (SMQ) Suicide and self-injury. |
|  | **Drug** | Any drugs coded as suspected or interacting |
| **Sampling** |  | Backward sampling, reviewing consecutive reports from the index date for a limited time box (2 working days, up to 151 reports). |
| **Relevance**<br>**Adjudication**<br>**(rule out**<br>**criteria)** | **Exposure** | See Table 1, no restriction on administration |
|  | **Event** | Events in which suicidality-related terms refer to the use of a drug as a method or context of suicidality, rather than to drug-induced suicidal ideation. In particular, overdose as an instrument of suicide/self-harm, abuse, misuse, medication error, or homicide. |
|  | **Relationship** | See Table 1 |
|  | **Population** | No restriction applied. Contextual features related to misuse, overdose, or non-therapeutic use could contribute to lack of relevance for event or relationship (e.g., overdoses as a means of self-injury). |

**Annotation was performed by one operator (MF) to evaluate the applicability of the framework** under routine-review conditions and quantify non-assessability. Patterns were compared across drugs and event terms to assess whether relevance varied systematically. Insights gained from this process informed further adjustments to the framework.

### 3.5 Data visualization

**Frequencies were used to summarize** the proportions of reports classified as relevant and non-assessable, and relevant reports that included explicit reporter suspicion. Visualization included Sankey plots (for streams of reports from retrieval to qualification), Upset plots (for the distribution of evidentiary qualifiers), and heat maps (for relevance patterns across drugs and event terms).

## 4. Results

### 4.1 Framework

**Across the two case studies, we developed a structured framework for case identification** that distinguishes retrieved reports from cases of interest through systematic adjudication of relevance. The framework evaluates each report across four domains (exposure, event, drug-event relationship, and setting/population), allows explicit classification of non-assessable cases when information is insufficient, and further qualifies relevant reports using reporter suspicion and evidentiary qualifiers. Applied in both information-rich and routine-review settings, the framework consistently identified substantial proportions of non-relevant reports and revealed distinct patterns of non-relevance across drugs and events, while providing a structured basis for interpreting the evidentiary value of individual reports.

**Figure 2.**
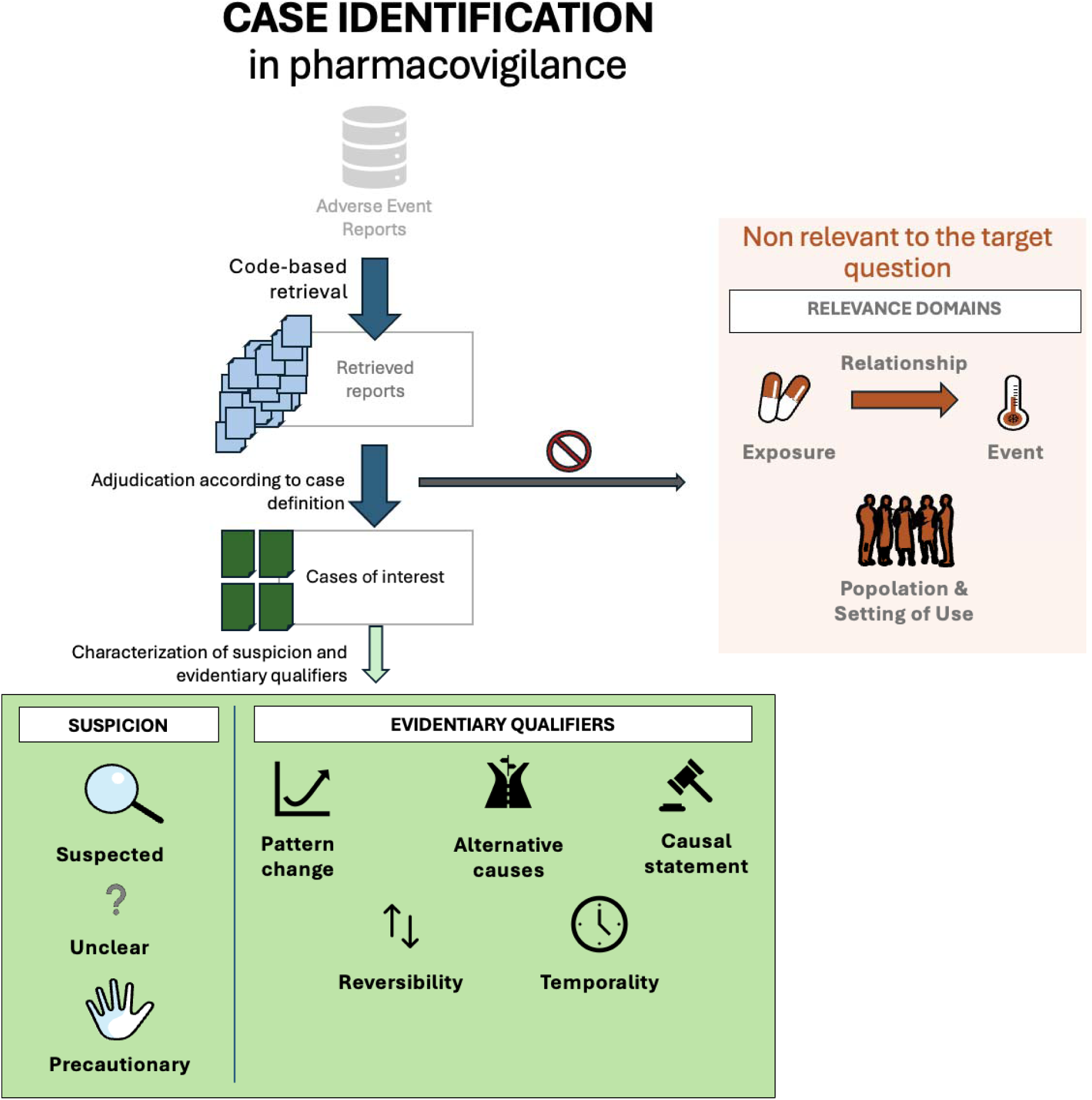
Case-identification framework for adjudication of retrieved reports to a case series.

### 4.2 Case Study 1 – Impulsivity and Medications

**From >36 million deduplicated reports in VigiBase** (27 Dec 2023), 40,722 were retrieved by the impulse-control disorders query [31]. After excluding reports without eligible narratives, candidate reports included 846 DAAs reports, 398 TGAs, 89 methylphenidate, 460 SSRIs, and 95 negative controls (Figure S1).

**Inter-annotator agreement was substantial** for both relevance (92% across 477 annotated reports–κ=0.74) and suspicion (93% across 375 annotated reports–κ=0.81). Disagreement on relevance was asymmetrical and mainly related to bystander situations; this was resolved by classifying definitive alternative attribution as non-relevant and co-suspected drugs as relevant with an alternative cause qualifier specification. Six reports were excluded due to unresolved disagreement (Table S1).

**Non-relevance varied by drug group**: lowest for positive controls (4% for DAAs and TGAs combined) and higher for methylphenidate (40%), SSRIs (20%), and negative controls (33%). Reasons for non-relevance also differed: event mismatch predominated for methylphenidate (non-reward-driven impulsivity in 60% of non-relevant reports), whereas relationship mismatch predominated for negative controls (bystander in 81% of non-relevant reports).

**Among relevant reports, explicit suspicion was more frequent for positive controls** (86%) than for uncertain (29%) or negative controls (33%) (Figure 3; Table S2). Reports with explicit suspicion more often included supportive qualifiers (e.g., causal statements, change in event pattern, compatible temporality, and reversibility; Figure 4, S2), precautionary reports more often included negative causal statements (Figure S3), and unclear reports typically lacked informative qualifiers (Figure S4).

**Figure 3.**
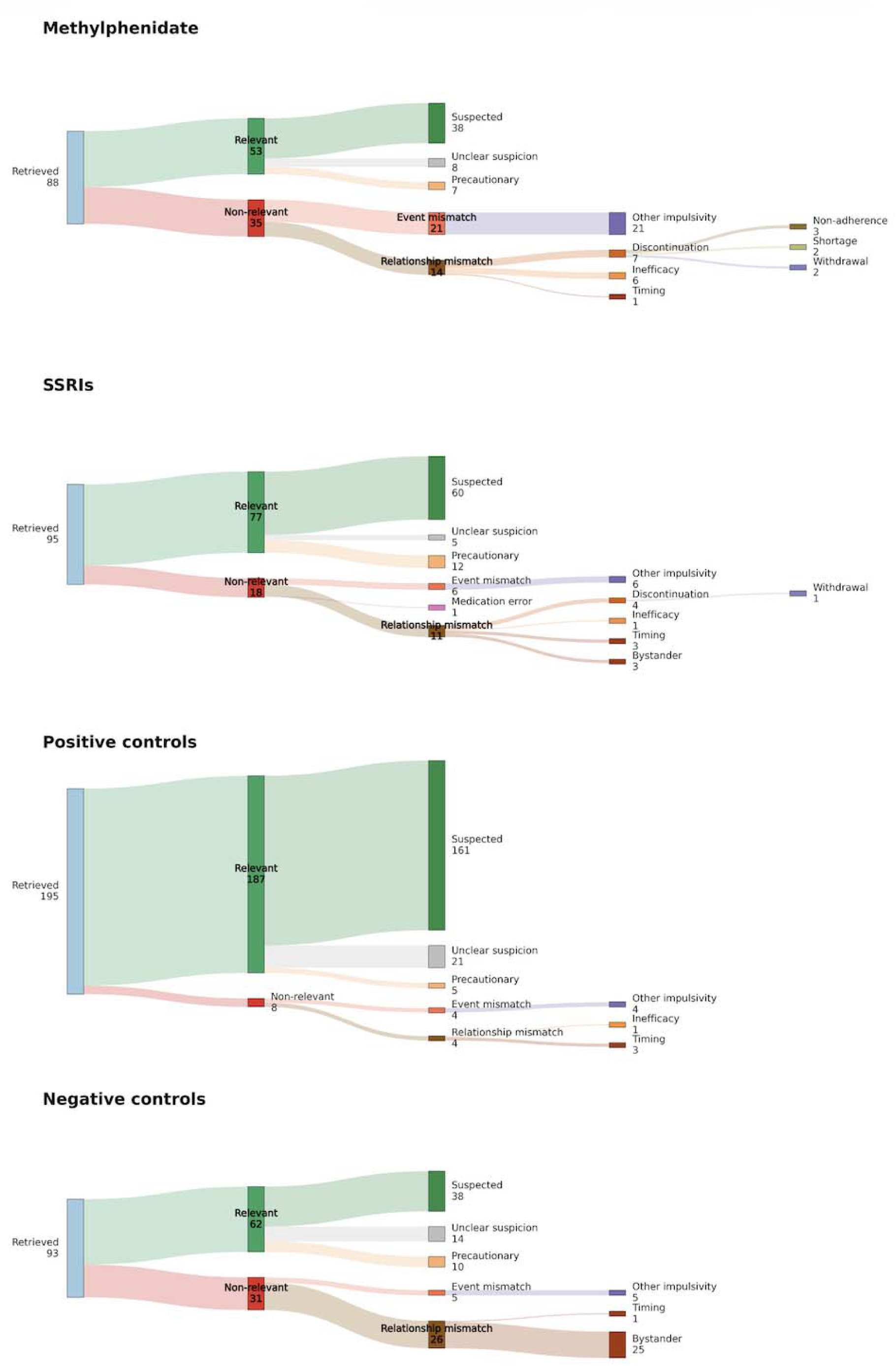
– Relevance of retrieved reports for evaluating impulsivity as a potential adverse reaction to methylphenidate and SSRIs, alongside positive and negative controls. Reports are processed from left to right: retrieved based on drug-event co-occurrence, adjudicated for relevance using the case definition, and characterized using evidentiary qualifiers of interest. This workflow enhances systematicity, transparency and interpretability of the case identification process.

**Figure 4.**
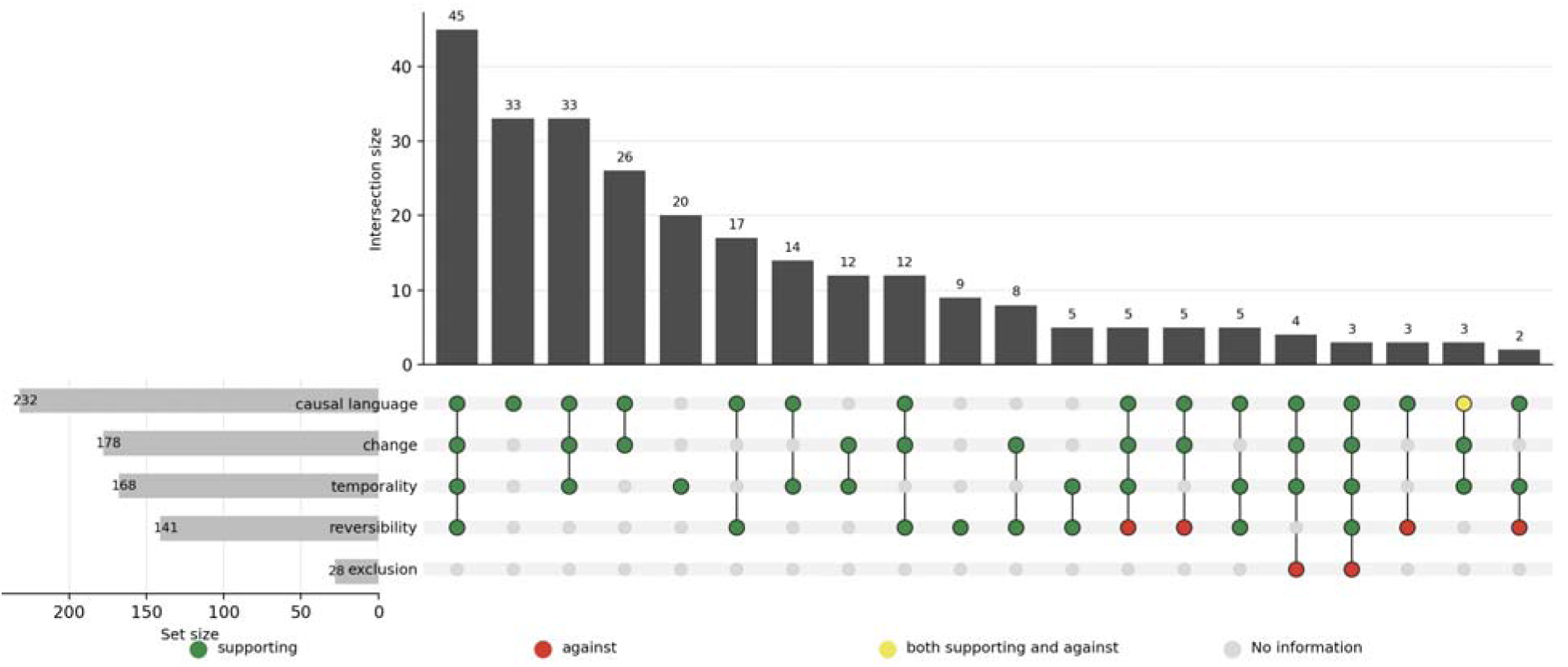
– most frequent (>5 cases) combinations of qualifiers in reports with explicit suspicion.

### 4.3 Case Study 2 – Suicidality and Medications

**From >43 million reports in VigiBase** (25 Jan 2026), 255,311 were retrieved by the narrow SMQ Suicide and self-injury. The annotated sample included 151 reports and 255 suspected drugs (129 unique), most commonly paracetamol (21), bupropion (19), and alprazolam (9). Frequent preferred terms were intentional overdose (50), suicidal ideation (41), and suicide attempt (34).

**Only 14% were relevant**, with 69% non-relevant and 17% non-assessable. Non-relevance was driven mainly by event mismatch (93%) as most reports reflected instrumental overdose rather than suicidality as an adverse reaction. Overdoses were heterogeneous in intent: most suicidal (84%) but also 3 homicidal administrations. Relationship mismatch (6%) included inefficacy, discontinuation, and bystander situations (Figure 5).

**Figure 5.**
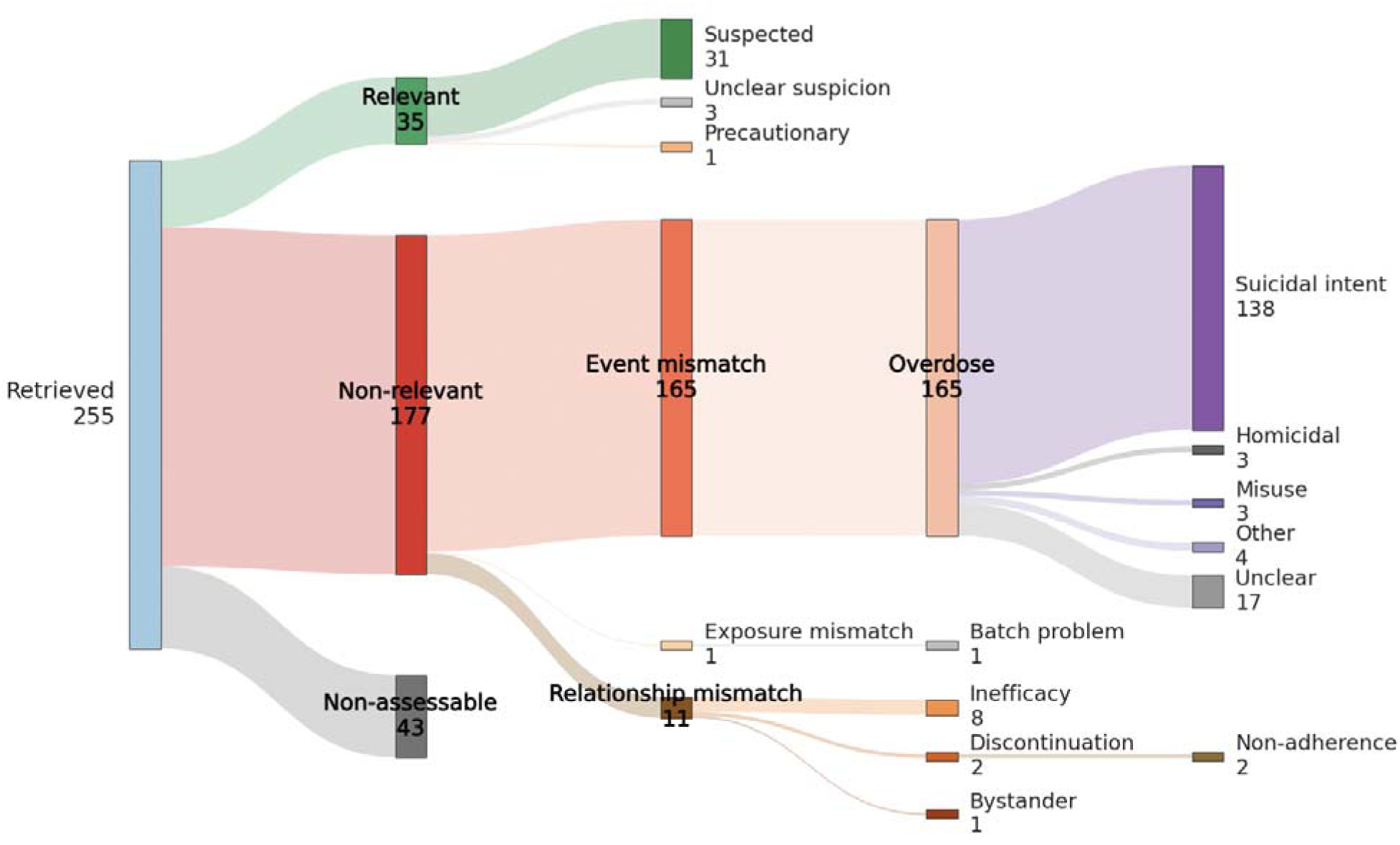
– Relevance of retrieved reports for evaluating suicidality as a potential adverse reaction to any active ingredient. Reports are processed from left to right: retrieved based on drug-event co-occurrence, adjudicated for relevance using the case definition, and characterized using evidentiary qualifiers of interest. This workflow enhances systematicity, transparency and interpretability of the case identification process.

**Relevance profiles varied markedly across drugs**. 78% of the 129 unique drugs had no relevant report. Some drugs showed higher proportions of relevant cases, though counts were small (e.g., semaglutide–3/4 reports, isotretinoin–4/6, venlafaxine–2/6) and should discourage any conclusion on the safety profile of these products. Frequently reported drugs (e.g., paracetamol, bupropion, alprazolam), were typically involved in overdose contexts and rarely relevant. Potassium was involved in reports of homicidal administration.

**Relevance profiles also varied by event term**. Four over nine documented suicidality terms had no relevant report (45%). Relevant reports were more common for suicidal behavior (5/6 reports) and ideation (26/41), whereas frequently retrieved terms (e.g., intentional overdose, suicide attempt, completed suicide) were largely non-relevant due to overdose contexts (Figures 6-7).

**Figure 6.**
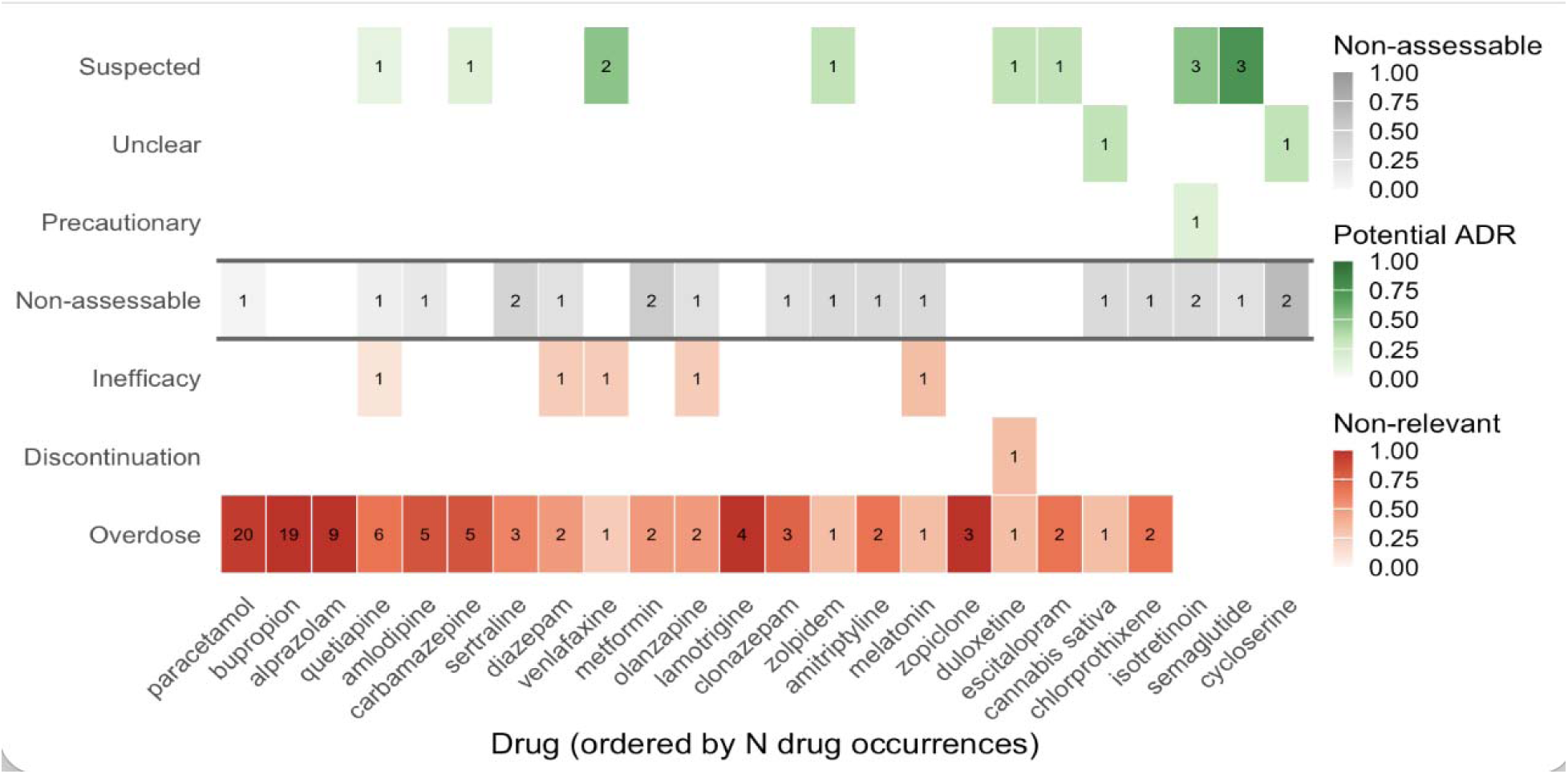
– Heatmap highlighting systematic misclassification patterns arising from code-based retrieval approaches for drugs with more than 2 occurrences, assessed in the context of suicidality as a potential adverse drug reaction. Drugs are displayed on the x-axis (ordered by frequency), and categories on the y-axis (grouped into non-relevant mechanisms, non-assessable reports, and potential adverse drug reactions). Cell color represents the proportion of reports within each drug assigned to a given category, while numbers indicate absolute counts. Given the small number of reports for many drugs, results should be interpreted descriptively rather than inferentially.

**Figure 7.**
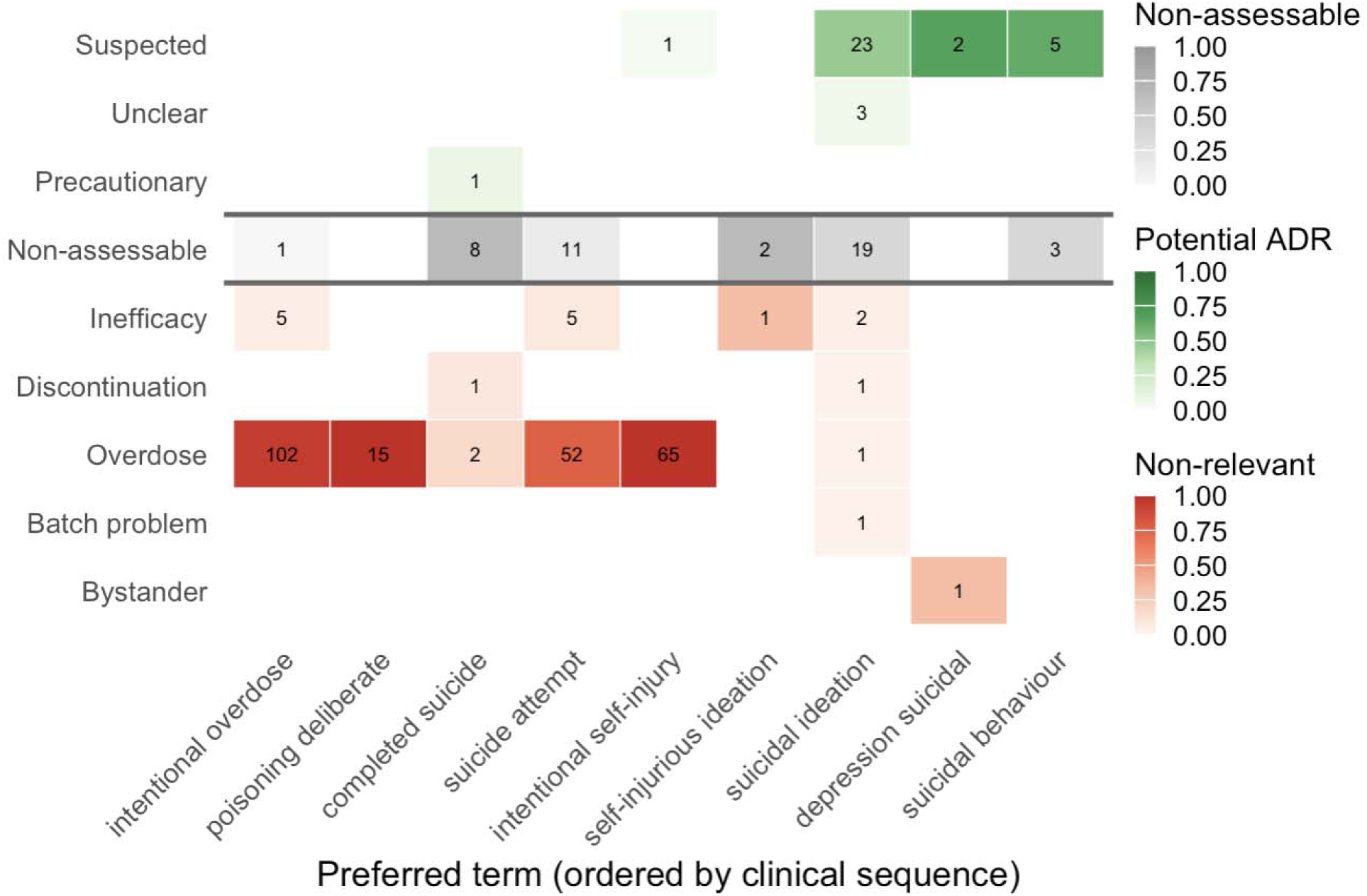
– Heatmap highlighting systematic misclassification patterns arising from code-based retrieval approaches for events (MedDRA preferred terms–PTs from the SMQ Suicide and Self Injury) with more than 2 occurrences, assessed in the context of suicidality as a potential adverse drug reaction. Events are displayed on the x-axis (ordered by frequency), and categories on the y-axis (grouped into non-relevant mechanisms, non-assessable reports, and potential adverse drug reactions). Cell color represents the proportion of reports within each PT assigned to a given category, while numbers indicate absolute counts. Given the small number of reports for many PTs, results should be interpreted descriptively rather than inferentially.

## 5. Discussion

### 5.1 Summary of main findings

**This paper should be read as a framework development study that intentionally includes two highly complex case studies to illustrate the challenges that may arise in case identification.** It should not be interpreted as an exhaustive signal management exercise for drug-induced impulsivity and suicidality. Across the two case studies, retrieval based on drug-event co-occurrence conflated clinically heterogeneous candidate reports that did not directly correspond to cases of interest, with potential to distort downstream inference. A substantial proportion of retrieved reports were non-relevant when assessed against explicit case definitions, with structured, question-specific patterns varying across drugs and events. Non-relevance was most pronounced for weakly defined or bias-prone safety questions, where structured coding captures clinical meaning less reliably. Non-assessability was pronounced in the information-constrained review setting.

### 5.2 Implications for case identification

**These findings reinforce that retrieval should be understood as generating candidate reports rather than validated cases**, due to diagnostic uncertainty, limited reporting context, miscoding, and the inability of structured coding to fully capture the clinical context. Case identification is therefore a central methodological operation for sound pharmacovigilance analysis, affecting validity, interpretability, and reproducibility. Case identification should be understood as beginning already at the retrieval stage, which should privilege sensitivity (so to not miss entirely relevant cases) and make use of shared, validated queries (see Table S3 for different queries used to retrieve suicidality-related reports). Case identification should then include transparent adjudication against predefined ruling out criteria, supported by domain expertise, and robust to unanticipated sources of non-relevance.

**The proposed framework structures relevance assessment on a-priori case definitions and clinical questions** across exposure, event, drug-event relationship, and context, and distinguishes relevance (whether a report should be included or not in a cases series) from reports qualifiers (what the specific contribution of the individual report within the case series is). Both dimensions are question-specific and should be adapted accordingly. A key strength of the framework is its ability to systematize the focus to few reports requiring detailed review. This may be especially valuable in time-sensitive settings, particularly if case identification and characterization can be automated and made more scalable through artificial intelligence. A key limitation of the framework is its dependence on the information available in individual reports, which may not be sufficient to clearly identify a report. As a result, non-assessability may be a more prominent issue in databases such as VigiBase than in national centres, where narrative information may be more readily available. Still, Case 2 on drug-induced suicidality showed that, beyond the issue of non-relevant reports, reports were more often judged to be non-assessable than relevant. In similar situations, the pipeline could indicate that further judgment based solely on adverse event reports should be suspended, and that other data sources may be needed to identify cases of interest less ambiguously.

**The framework may also support exploratory signal detection** by organizing heterogeneous retrieval outputs, distinguishing clinically distinct report types, and better defining the generated hypothesis. For example, the framework can help derive, from a candidate set prioritized by a signal of disproportionate reporting, a hypothesis of adverse drug reaction not only concerning an active ingredient and an event term, but for example, a specific administration route, in a setting of recreational use among young individuals, and resulting in a clinical manifestation with certain characteristics. Case identification as report adjudication according to a case definition and case definition refinement based on retrieved reports can be integrated in a series of iterations.

### 5.3 Interpreting non-relevant reports

**Non-relevance should not be treated as noise.** Relevance is question-dependent and not an intrinsic property of reports, and many reports non-relevant to adverse drug reactions reflect meaningful medicine-related problems (e.g., lack of effect, misuse, overdose, medication errors, shortage, non-compliance) that warrant explicit characterization rather than residual classification.

**The proposed qualifier framework, oriented toward adverse drug reactions, may require adaptation for other contexts**, where different dimensions (e.g., intent and dose for overdose, process-level and root-cause for medication errors, personal motivation for non-adherence) are more informative.

### 5.4 Limitations

**The framework was developed in two case studies and not formally validated**; its generalizability and impact in signal management remain to be established. Sample sizes were modest, and findings illustrate patterns rather than prevalence or correcting factors [36], which were outside the objectives of this study. Case 2 was restricted to recent reports, which may not capture longer-term reporting dynamics (e.g., shifts due to clinical awareness, reporting behavior, coding practice, and regulatory attention) and may include clustered reports (e.g., originated in the same lawsuit).

The two case studies were selected precisely because of their complexity. For questions like whether methylphenidate can paradoxically cause impulsivity and whether a drug can cause suicidal intention, adverse event reports, especially when free text narratives are unavailable, may not be the most suitable data source. This reflects several challenges: the absence of unambiguous diagnostic criteria, unlike for conditions like drug-induced liver injury [10]; substantial reverse causality, since these drugs may be used on- or off-label to treat the reported condition; and the strong influence of regulatory warning and reporter type (and whether they have an interest or obligation to report, which can sometimes put into question the clinical meaning of the event; see Figure S5).

**Adjudication relied on incomplete information and involved interpretive judgement**, with residual subjectivity despite substantial agreement. Case 2 used a single annotator.

**Blinding to drug identity was not feasible**, and prior expectations may have influenced annotation.

### 5.5 Future directions

**Future work should focus on scalable workflows** combining structured rules, expert review, and automation. Artificial intelligence may assist in identifying relevant reports and extracting contextual features but requires careful evaluation.

**Variation in non-relevance across drugs and events, suggests value in targeted validation efforts**. For some questions, retrieval output is more heterogeneous, and broader data access (e.g., beyond structured text as publicly available in FAERS) may be needed to avoid misinterpretation. This could well be the case for complex outcomes like suicidality or paradoxical reactions.

**Further work is needed to define relevance across drug-event questions** and to improve reproducibility through reusable and auditable case definitions, with implications for database design, reporting practices, data processing pipelines, and integration with other qualifiers (e.g., completeness [25,26], evidentiary strength [27,28], reporter suspicion [29]).

## 6. Conclusion

**Drug-event co-occurrence is a powerful retrieval strategy, but it is not case identification.** It generates a candidate set, not a validated case series. Determining whether a retrieved report is truly relevant requires explicit, structured assessment of exposure, event, context, and drug-event relationship.

**Case identification should therefore be treated as a core methodological step.** Transparent and reproducible case-definitions, combined with systematic adjudication, are essential for producing interpretable and reliable pharmacovigilance analyses. The framework proposed here offers a practical way to structure and document this process, while remaining adaptable to the specific clinical question. As appears to be the case here for drug-induced suicidality, it may sometimes highlight substantial challenges in unambiguously identifying cases of interest and therefore underscore the need to use other data sources.

**Reports classified as non-relevant should not be dismissed as noise.** They often reflect other meaningful medicine-related problems, such as lack of effect, misuse, medication error, that warrant explicit consideration in their own right.

**Ultimately, improving reliability of pharmacovigilance results** does not depend only on retrieving more data, but on better distinguishing which data truly answer the question being asked.

## Statements and Declarations

### Funding

No funding was received for conducting this study.

### Competing interests

None to declare

### Ethics approval

Not applicable. The data in VigiBase is de-identified and not considered personal data.

### Consent to participate

Not applicable. This study did not involve human participants.

### Consent to publish

Not applicable.

### Authors’ contributions

All authors participated in the conceptualization and design of the study. MF led the development of the draft with contributions from JF, DS, VG, NN. MF, JF, DS, VG, FvH, JS, LH, NN, JE participated in focus groups to conceptualize the framework. MF and JF performed the annotation. MF performed the data analysis. All authors contributed to the review of the manuscript. All authors have read and approved the final version.

### Availability of data and materials

The data that support the findings of this study are not publicly available. Access to data in VigiBase is subject to the requirements of the VigiBase Data Access Conditions. For the purposes of this study, access to the data was granted following case-by-case review in accordance with those Conditions (IR 00122698). Subject to these conditions, data is available on reasonable request. For further inquiries, please contact Uppsala Monitoring Centre via https://who-umc.org/contact-information

https://github.com/PVverse/caseid-paper

### Code availability

The code used for this study is made available on https://pvverse.github.io/caseid-paper/. For further inquiries, please contact Uppsala Monitoring Centre via https://who-umc.org/contact-information/.

## Acknowledgment

The authors are indebted to the members of the WHO Programme for International Drug Monitoring who contribute reports to VigiBase. However, the opinions and conclusions of this study are not necessarily those of Uppsala Monitoring Centre, LAREB, the Drug Safety Research Unit, the University of Gröningen, the various member organisations, nor the WHO.

## Footnotes

1 Potentially using hierarchical terminologies, Standardized MedDRA Queries, MedDRA Labeling Groupings, and custom query definitions [4–6]

2 Requiring coding of all reported information regardless of causal association. As a consequence, correctly coded reports may still be off target for a specific analytic question.

## Notes

### Competing Interest Statement

The authors have declared no competing interest.

### Author Declarations

Ethical board review was not required for this study, as this study did not use personal data. Data was used according to Data Access Conditions from the VigiBase WHO database of adverse event reports. For the purposes of this study, access to the data was granted following case-by-case review in accordance with those Conditions (IR 00122698).

