## Supplementary Material for "Towards a Framework for Case Identification in Pharmacovigilance: Not All Reports are Created Equal"

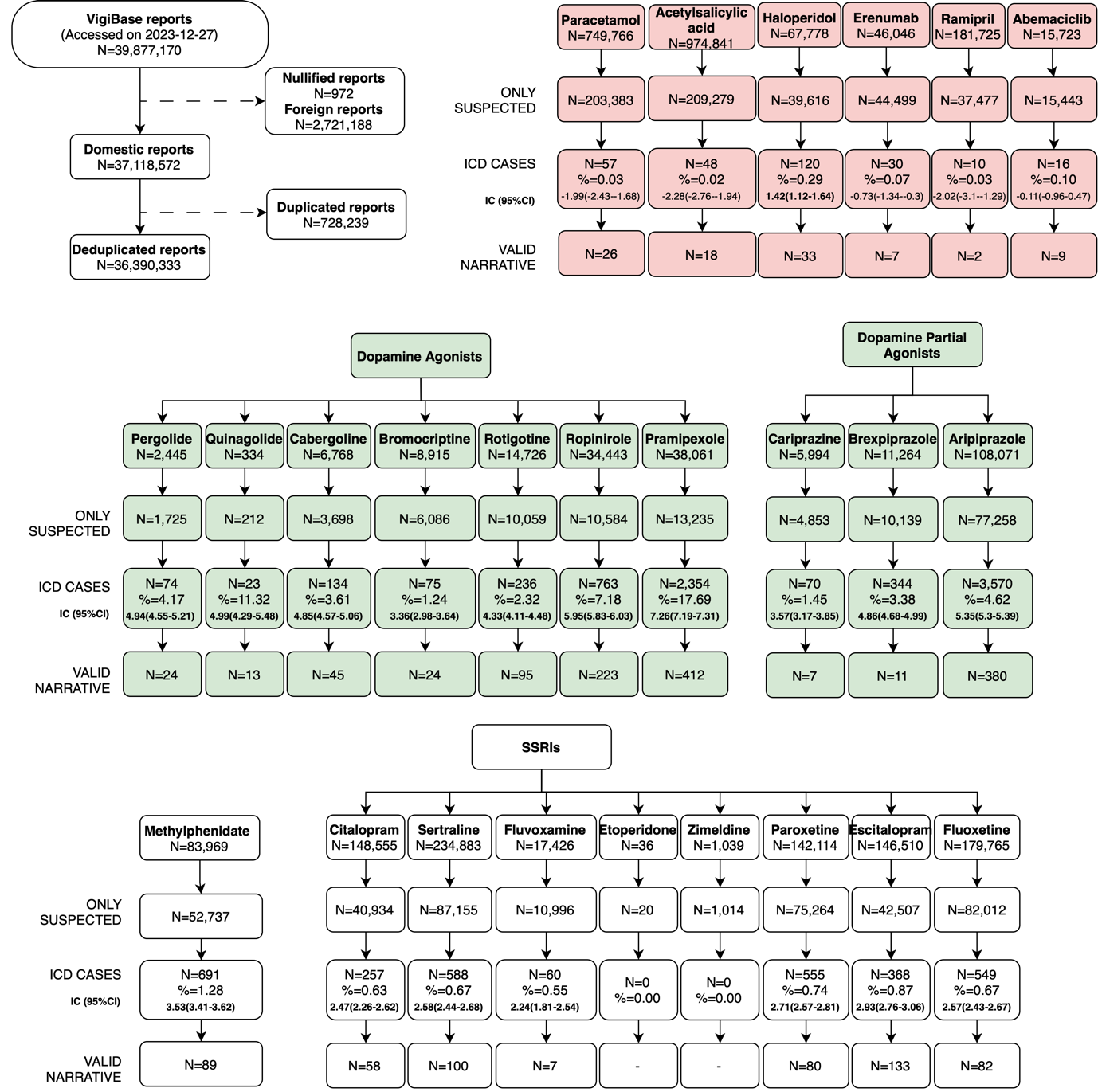

Figure S1: flowchart showing the outcome of the case retrieval of Case Study 1.

Table S1: disagreement documentation Case 1

| **Task** | **N** | **% agreement** | **K Cohen** | **mcNemar** | **Reasons (JF vs MF)** |
| --- | --- | --- | --- | --- | --- |
| Relevance | 477 | 91.82  =>  98.95 | 0.739  =>  0.970 | 1.917*10^-5^  =>  0.074 | 31 reaction vs bystander;  2 inefficacy vs reaction;  2 non-adherence vs reaction;  1 other event vs reaction;  1 reaction vs inefficacy;  1 impossible timing vs discontinuation;  1 relationship unk vs non-adherence  =>  5 reaction vs bystander |
| Suspicion | 375 | 92.80  =>  99.74 | 0.814  =>  0.993 | 0.327  =>  1.000 | 10 precautionary vs suspected  6 unclear vs suspected  6 suspected vs unclear  4 suspected vs precautionary  1 precautionary vs unclear  =>  1 unclear vs suspected |

Table S2: annotation results Case 1

| **Group** | **Drug** | **Irrelevant %** | **Irrelevance Reason** | **Relevant %** | **Relevant suspicion** | **Median [IQR] completeness** |
| --- | --- | --- | --- | --- | --- | --- |
| Positive | DAA | 2 (2.04%) | 1 other impulsivity  1 timing | 96 (97.96%) | 81 (84.38%) suspected  3 (3.13%) precautionary  12 (12.5%) unclear suspicion | 0.60 [0.35-0.90]  0.90 [0.90-0.93]  0.35 [0.15-0.50] |
| Positive | TGA | 6 (6.19%) | 2 timing  1 inefficacy  3 other impulsivity | 91 (93.81%) | 80 (87.91%) suspected  2 (2.17%) precautionary  9 (9.89%) unclear suspicion | 0.63 [0.36-0.90]  0.95 [0.93-0.98]  0.40 [0.39 vs 0.90] |
| Ambiguous | Methylphenidate | 35 (39.77%) | 21 other impulsivity  6 inefficacy  2 withdrawal  3 non-adherence  2 drug shortage  1 timing | 53 (60.23%) | 38 (71.7%) suspected  7 (13.21%) precautionary  8 (15.09%) unclear suspicion | 0.70 [0.46-0.90]  0.54 [0.47-0.63]  0.25 [0.15-0.90] |
| Ambiguous | SSRI | 18 (19.95%) | 6 other impulsivity  3 inefficacy  3 bystander  1 withdrawal  4 timing  1 medication | 77 (81.05%) | 60 (77.92%) suspected  12 (15.58%) precautionary  5 (6.49%) unclear suspicion | 0.67 [0.46-1.00]  0.45 [0.33-0.83]  0.32 [0.32 vs 0.35] |
| Negative | All drugs together | 31 (33.23%) | 25 bystander  5 other impulsivity  1 timing | 62 (66.67%) | 38 (62.29%) suspected  10 (16.13%) precautionary  14 (22.58%) unclear suspicion | 0.49 [0.39-0.96]  0.48 [0.35-0.63]  0.57 [0.42-0.69] |

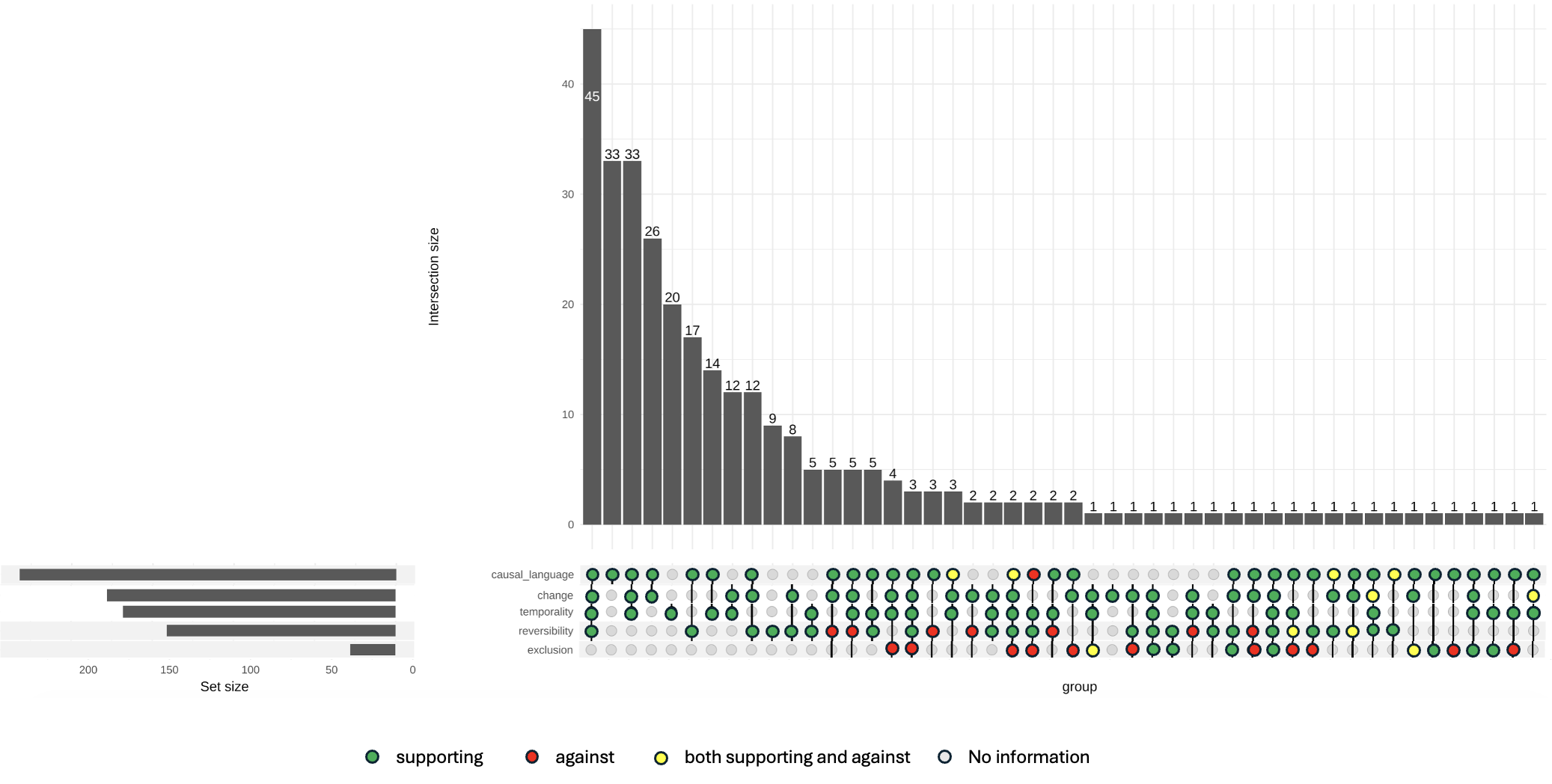

Figure S2: evidentiary indicators for suspected reports

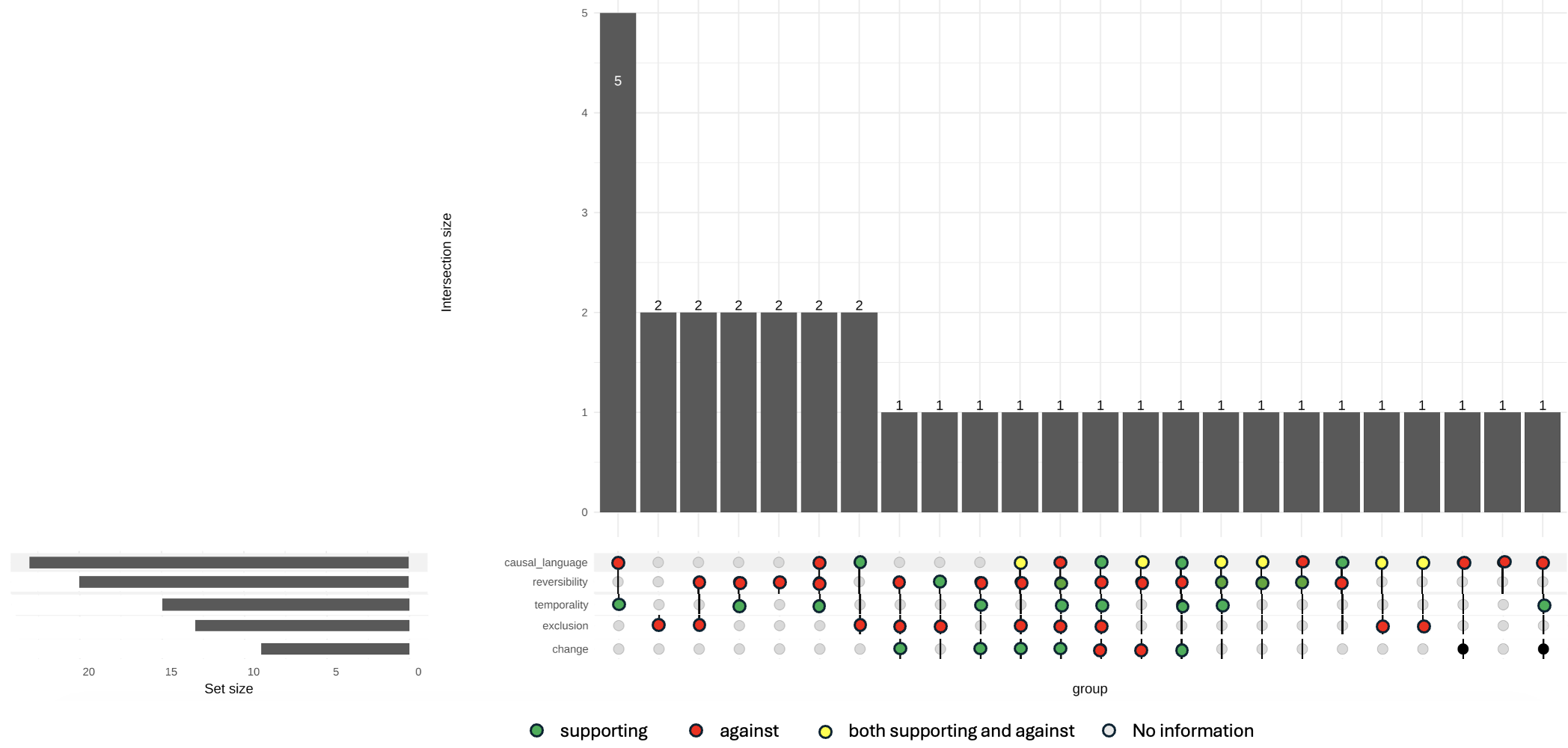

Figure S3: evidentiary indicators for precautionary reports

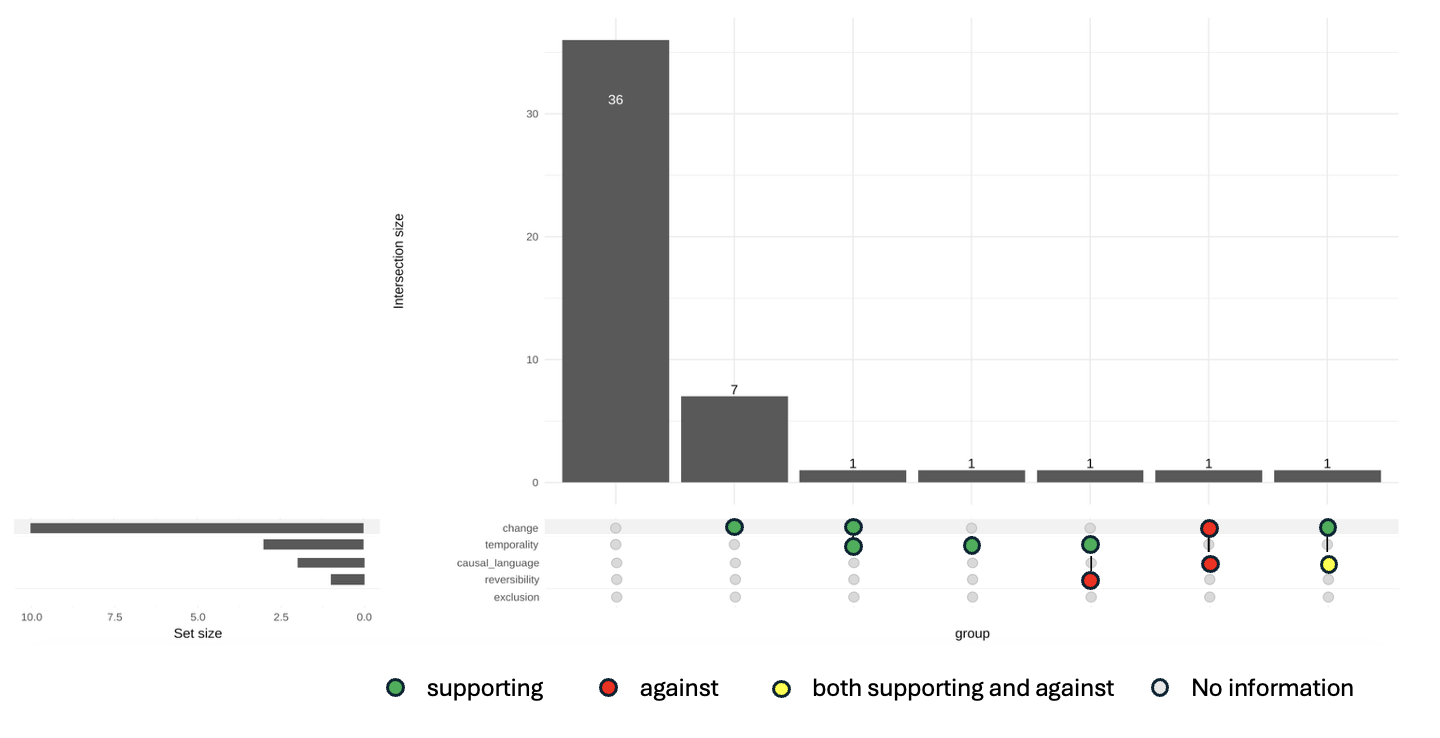

Figure S4: evidentiary indicators for reports with unclear intention

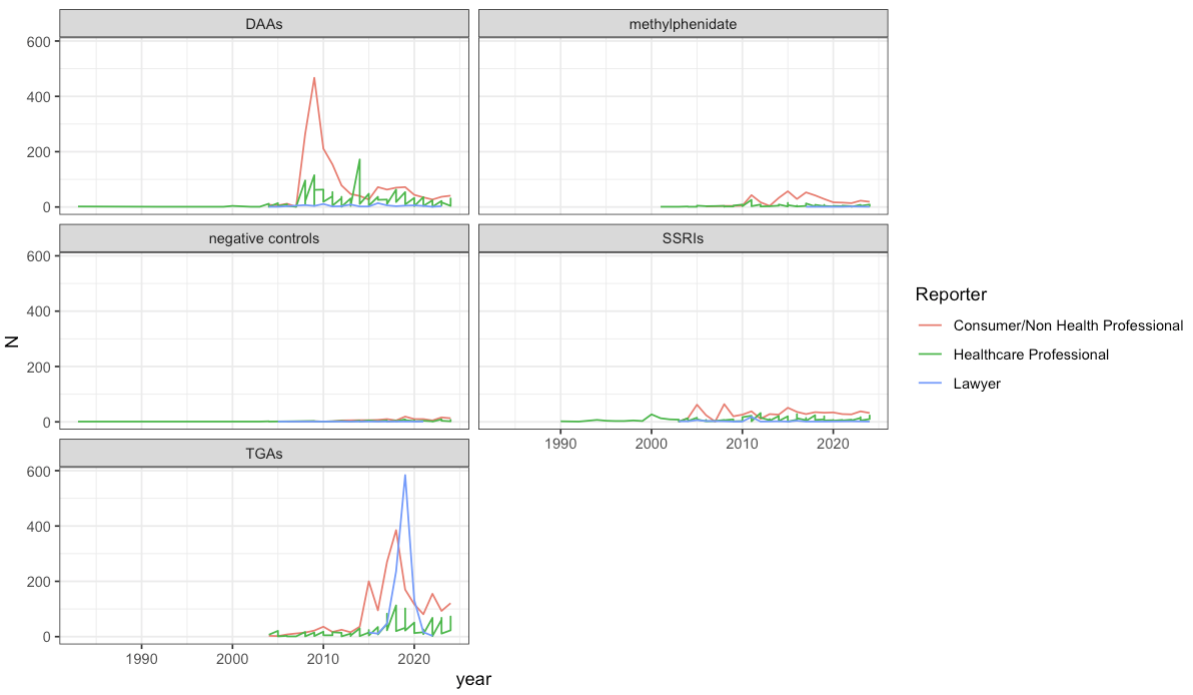

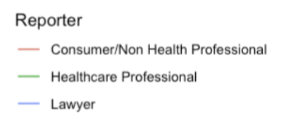

Figure S5: temporal and reporter trend for drug-induced impulsivity, separated by drug class.

Table S3: definitions used by studies investigating suicide in adverse event reports published in the last two years (number referring to bibliography below). Green cells imply the PT was included for the retrieval and is included in the SMQ suicidal and self-injurious behavior (Narrow). Red cells imply the PT was included for the retrieval and is not included in the SMQ.

|  | 1 | 3 | 5 | 6 | 8 | 2 | 4 | 9 | 10 | 11* | 12 | 13 | 7** |
| --- | --- | --- | --- | --- | --- | --- | --- | --- | --- | --- | --- | --- | --- |
| assisted suicide |  |  |  |  |  |  |  |  |  |  |  |  |  |
| columbia suicide severity rating scale abnormal |  |  |  |  |  |  |  |  |  |  |  |  |  |
| completed suicide |  |  |  |  |  |  |  |  |  |  |  |  |  |
| depression suicidal |  |  |  |  |  |  |  |  |  |  |  |  |  |
| intentional overdose |  |  |  |  |  |  |  |  |  |  |  |  |  |
| intentional self-injury |  |  |  |  |  |  |  |  |  |  |  |  |  |
| poisoning deliberate |  |  |  |  |  |  |  |  |  |  |  |  |  |
| self-injurious ideation |  |  |  |  |  |  |  |  |  |  |  |  |  |
| suicidal behaviour |  |  |  |  |  |  |  |  |  |  |  |  |  |
| suicidal ideation |  |  |  |  |  |  |  |  |  |  |  |  |  |
| suicide attempt |  |  |  |  |  |  |  |  |  |  |  |  |  |
| suicide threat |  |  |  |  |  |  |  |  |  |  |  |  |  |
| suspected suicide |  |  |  |  |  |  |  |  |  |  |  |  |  |
| suspected suicide attempt |  |  |  |  |  |  |  |  |  |  |  |  |  |
| suicide of companion |  |  |  |  |  |  |  |  |  |  |  |  |  |
| columbia suicide severity rating scale |  |  |  |  |  |  |  |  |  |  |  |  |  |

*based on presence of “suic” in the PT.

**Full paper not available

### Appendix A – Query for retrieval of reward-driven impulsivity

The PT used for the retrieval were:

- 'Gambling', 'Gambling disorder',
- 'Compulsive shopping',
- 'Hyperphagia', 'Food craving', 'Binge eating', 'Increased appetite'
- 'Erotophonophilia', 'Frotteurism', 'Excessive masturbation', 'Excessive sexual fantasies', 'Exhibitionism', 'Fetishism', 'Compulsive sexual behaviour', 'Hypersexuality', 'Kluver-Bucy syndrome', 'Libido increased', 'Masochism', 'Paedophilia', 'Paraphilia', 'Sadism', 'Sexual activity increased', 'Sexually inappropriate behaviour', 'Transvestism', 'Voyeurism',
- 'Compulsive hoarding',
- 'Excessive exercise',
- 'Gaming disorder',
- 'Kleptomania', 'Shoplifting',
- 'Overwork',
- 'Poriomania',
- 'Pyromania',
- 'Impulse-control disorder', 'Impulsive behaviour', 'Behavioural addiction', 'Disinhibition'

#### Appendix B – Annotation decision tree

##### Annotation Unit

Annotation of each report at the drug-event level, considering only suspected and interacting drugs, so that even when investing a specific event, the same report can contain multiple drugs with different functional roles (beyond them being reported as suspected).  Each drug could potentially have multiple roles for the same event.

#### Input

All available information in the report, including both coded fields and free text, including coded events (MedDRA Preferred Terms (v26.1)), drugs (active ingredients standardized through WHODrug), free-text narratives (ICH field “Case Narrative Including Clinical Course, Therapeutic Measures, Outcome and Additional Relevant Information” (H.1)), temporal relationships, medical history (structured/free text fields from “Relevant Medical History and Concurrent Conditions”, D7), reporter causality assessment (“Result of Assessment” field (G.k.9.i.2.r.3), standardized to likely, possible, unlikely), dechallenge/rechallenge. Narratives in non-English languages were translated using Meta NLLB-200, run on internal servers for privacy compliance.

#### Assessing Relevance

Decide whether this drug-event pair within the report is relevant to the specific pharmacovigilance question. The relevance of a drug-event pair within a report depends on **relevance** of:

- *exposure*: marketed products or falsified drug, irrelevant dosage or route of administration, miscoding…
- *event*: respecting or not diagnostic criteria:
  - For suicide, we considered irrelevant events the **overdoses** of the drug as a means. Also look at the setting for the intention of the overdose.
  - For reward-driven impulsivity, we considered irrelevant events referring to compulsions, aggressivity, and suicidality.
- *relationship*: referring or not to the relationship of interest. Different relationships between drug and event are:
  - **reaction:** The drug is framed as causally contributing to the event (e.g., causing impulsivity or suicidal ideation). This relationship could actually have different suspicion by the reporter:
    - **suspected**
    - **precautionary** (with explicit strong doubt that the event results from the exposure)
    - **unclear**
  - **bystander**: the event is explicitly attributed to another drug.
  - **inefficacy:** Phenomenon framed as relapse/worsening due to inefficacy or non-response while on treatment.
  - **discontinuation:** Phenomenon follows stopping, missing, or inability to access drug, due to shortage, withdrawal, or noncompliance; narrative suggests rebound or withdrawal context.
  - **impossible timing**: the event occurred before the exposure
- *setting*:
  - **self-**harm/suicidal act
  - **self-**treatment (misuse)
  - **performance**-enhancement
  - **recreational use** (abuse)
  - **poisoning** of someone else
  - **medication error**
  - **unclear** setting/intent
  - **Population with a specific age/sex**
- **unclear** relationship

#### Flagging Evidentiary Indicators

Information included in reports can constitute stronger or lower evidentiary support and opportunities for characterization. Here we focused on the evidentiary indicators for suspected adverse drug reactions, but information of interest to other medicine related problems could be different (e.g., who performed the medication error, what was the root cause of the medication error, what was the intention underlying the overdose).

1. **Temporality** (onset after start/dose change; latency plausible)
2. **Symptom change** (worsening/improvement described)
3. **Reversibility**: dechallenge (e.g., abated after stopping), rechallenge (recurrence after restart, if present), or dose-relation.
4. **Causal language** (explicit attribution)
5. **Alternative explanations considered/excluded** (e.g., role of the underlying disease or of a concomitant medication)
